## Supplementary materials for "Quantitatively assessing early detection strategies for mitigating COVID-19 and future pandemics"

**Supplementary Text**

Earliness of lockdown versus lockdown success for 85 countries’ first 2020 COVID-19 lockdowns.

To test the importance of early response, we asked whether countries that started COVID-19 lockdown earlier were better able to achieve low case counts post-lockdown (Fig. S1). We gathered (1) country lockdown dates from media reports (table S1), and combined these with (2) country-level COVID-19 confirmed case counts^73^ to estimate the infectious number of cases in each country at the time of lockdown. We gathered complete data for 85 countries and only analyzed these countries. In 2020, countries which instituted lockdown before 1,000 infectious cases were much more likely to contain COVID-19 initially, and this earliness of lockdown was more predictive of lockdown success than lockdown duration (Fig. S1). A lockdown was deemed successful if the average number of daily cases in the 7 days following lockdown is fewer than 10 cases. Of the 85 countries, 68 started lockdown before 1,000 infectious cases. 38% (26/68) of these countries with earlier lockdowns contained COVID-19 initially, compared to 0% (0/17) of countries with later lockdowns (a statistically significant difference at p = 0.0057). This is robust across many thresholds and definitions of lockdown success (Fig. S4) and caseload-based definitions of lockdown earliness (Fig. S5). Earliness of lockdown (measured by caseload) was also more predictive of lockdown success than geographical location (Fig. S2) and earliness measured by the raw lockdown start date (Fig. S3).

This analysis has limitations. First, we do not account for significant variation among countries in the extent of COVID-19 testing, the number of imported cases (approximated by the amount of travel), demographics and age structure, country size and density, and other factors; all of these can affect measurements of lockdown success. Countries which did not test extensively may be recorded as having low cases post-lockdown and as having started lockdown early, which could partially explain the observed association between lockdown earliness and success. However, this does not seem to explain most of the relationship: countries like Thailand and New Zealand, which tested extensively per capita^74^, were among the countries with early, successful lockdowns. Second, we cannot definitively distinguish correlation from causation: the association between earlier lockdown and fewer cases post-lockdown may be because countries which were willing to implement lockdown earlier were also more willing to implement and comply with more stringent lockdowns. Nevertheless, the observed association between lockdown earliness and success is consistent with the hypothesis that lockdown earliness improves chances of success.

We used (1) and (2) to calculate (a) the infectious number of cases in each country at the time of lockdown (as a measure of the earliness of lockdown) and (b) the number of cases in each country following lockdown (as a measure of the lockdown’s success). For (1), we only analyzed countries’ first lockdowns and required these lockdowns to start before 2021-01-01 and last longer than 7 days. When a country had regional lockdowns which differed from the national lockdown, we used the start and end dates of the national lockdown. If the country had no national lockdown and only regional lockdowns, we used the dates of the regional lockdown with the median start date.

For (a), we used overall case counts to estimate the *infectious* cases at the lockdown start date by assuming (i) a 5-day infectious period^47^ and therefore (ii) that the infectious cases on day T are 1/5 * (cases on day T-4) + 2/5 * (cases on day T-3) + … + 5/5 * (cases on day T). We also performed a sensitivity analysis using the raw case count average at the start of lockdown instead of the infectious case count (Fig. S5). For (b), we calculated the average daily new cases for the 7 days after the lockdown was lifted, and we considered a lockdown successful if this average was fewer than a threshold of 10 cases. We performed a sensitivity analysis for thresholds of 3, 10, 30 and 100 cases (Fig. S4).

To calculate statistical significance of the different lockdown success rates between countries with earlier and later lockdowns, we used the 2-sample test for equality of proportions implemented in R’s prop.test.

Validation of model in US states.

In validating our model in US state data, we were able to predict US state detection times with a mean absolute error of 0.97 weeks (Fig. S7). We achieved this with a relatively simple validation setup: R0 was the only parameter we allowed to vary among states, which did not allow the model to account for differing state testing turnaround and capacity or inter-state variation in growth rate of imported cases. Gathering those data and accounting for those could improve the model’s accuracy. (Other inter-state variables like differing age structures and demographics, as well as lockdown policies and pandemic-induced mobility changes, should be accounted for in the state-specific R0’s.)

Derivation for mathematical approximation of cases until detection

As an intuitive summary of the derivation, we break down the number of cases until detection into two variables: (i) the cases that occur until the infection of the “threshold case” (the final case needed to trigger detection), and (ii) the cases that occur afterwards during the delay between the threshold case’s infection and detection. In the formula, $d/p_{test}$ corresponds to (i), and each of those $\left( R_{0}-1 \right)/R_{0}$ * $d/p_{test}$ cases in the last generation spawns an outbreak process proportional to $\sum_{n=1}^{g} R_{0}^{n}$ cases, corresponding to (ii).

Full derivation: Assume the outbreak starts in a community covered by the detection system. We want the mean and variance of the cumulative number of cases $C$ which have occurred by the time the detection system is triggered. The outbreak occurs in generations, where the index case is generation 0 and each generation of cases creates the next generation of cases. We can express $C$ as follows:

$$C=T+\sum_{n=1}^{g} C_{n}$$

where $T$ is the number of cases infected until the threshold case is infected, $C_{i}$ is the number of infectious cases in the $i$-th generation after the threshold case is infected, and $g$ is the number of generations which occur in the delay between the threshold case’s infection and detection. Note first that

$$T\sim NBinom\left( d,p_{test} \right)$$

where $d$ is the detection threshold (the number of cases which need to be detected to constitute an outbreak) and $p_{test}$ is the probability any particular case is tested. For example, in the hospital system, $p_{test}=hosp\_rate$.

For the mean of $C$, we will need:

$$\mathbb{E}\left[ C_{n} \right]=\mathbb{E}\left[ \mathbb{E}\left[ C_{n}|T \right] \right]\approx\mathbb{E}\left[ \frac{R_{0}-1}{R_{0}}TR_{0}^{n} \right]=\left( R_{0}-1 \right)\left( R_{0}^{n-1} \right)\frac{d}{p_{test}}$$

The first expansion of $\mathbb{E}\left[ C_{n} \right]$ derives from two facts: (a) $C_{n}$ is the sum of approximately $\frac{R_{0}-1}{R_{0}}T$ independent and identically distributed branching processes, so that the mean of $C_{n}$ is $\frac{R_{0}-1}{R_{0}}T$ times the mean of one branching process. (b) From branching process mathematics, the mean of $Z_{n}$, the number of entities in the $n$-th generation of a branching process, is $\mathbb{E}\left[ O \right]^{n}$^75^, where $O$ is the offspring distribution (the distribution of the number of secondary cases infected by each primary case). In this study, $O$ is negative binomial with mean $R_{0}$ and dispersion 0.01^52^. Thus,

$$\mathbb{E}\left[ C \right]=\mathbb{E}\left[ T \right]+\sum_{n=1}^{g} \mathbb{E}\left[ C_{n} \right]\approx\frac{d}{p_{test}}\left( 1+\left( R_{0}-1 \right)\sum_{n=1}^{g} R_{0}^{n-1} \right)=\frac{d\times R_{0}^{g}}{p_{test}}$$

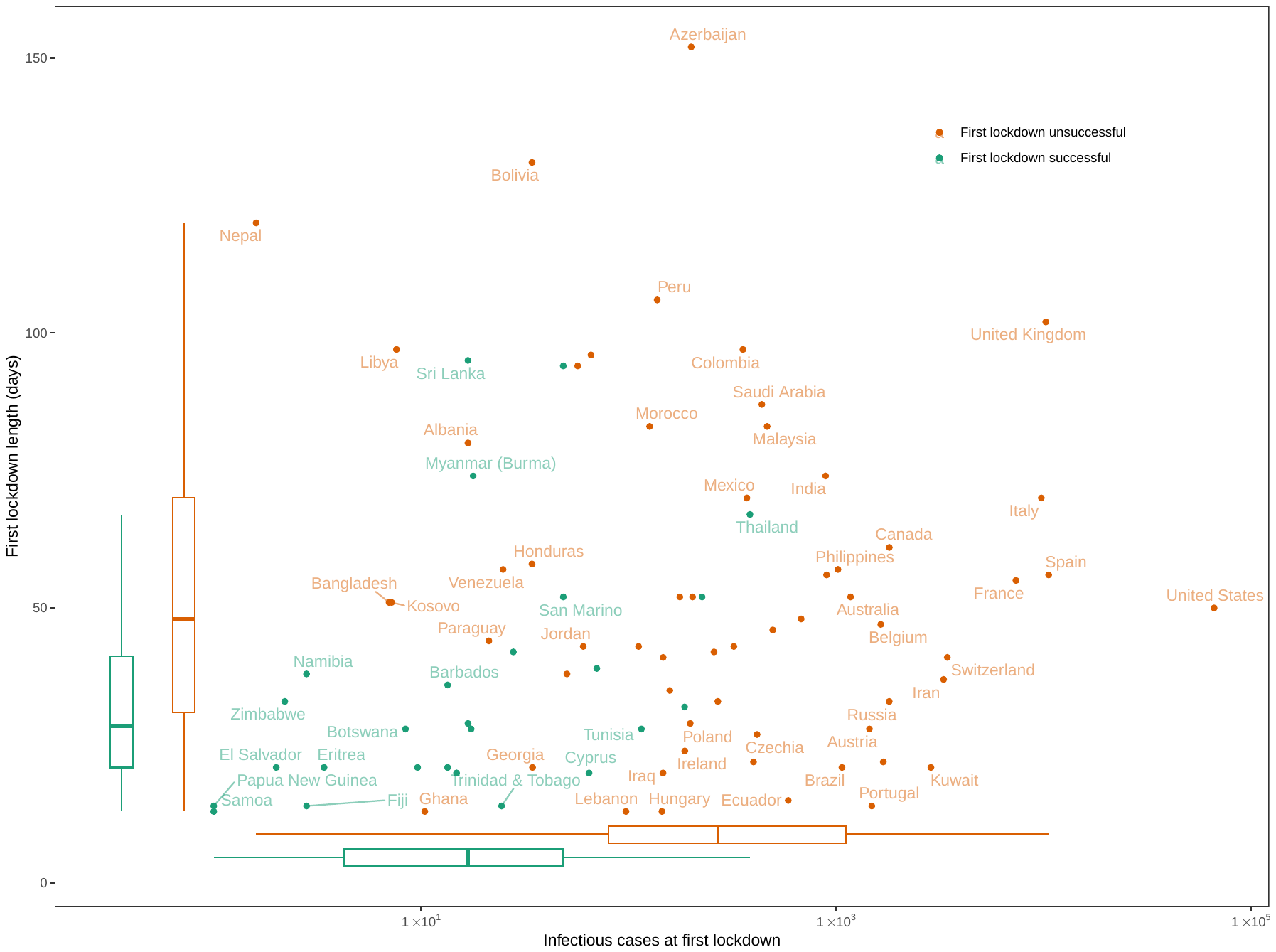

**Fig. S1.** Earliness of lockdown (x-axis) versus lockdown length in days (y-axis) and lockdown success for 85 countries’ first 2020 COVID-19 lockdowns (first lockdown unsuccessful (orange) and first lockdown successful (teal)). Earliness is measured by the number of infectious cases at the start of lockdown. A lockdown is successful if the average number of daily cases following lockdown is less than 10 for 7 days. Boxplots on the axes show marginal distributions and the separation of successful and unsuccessful lockdowns by lockdown earliness and length.

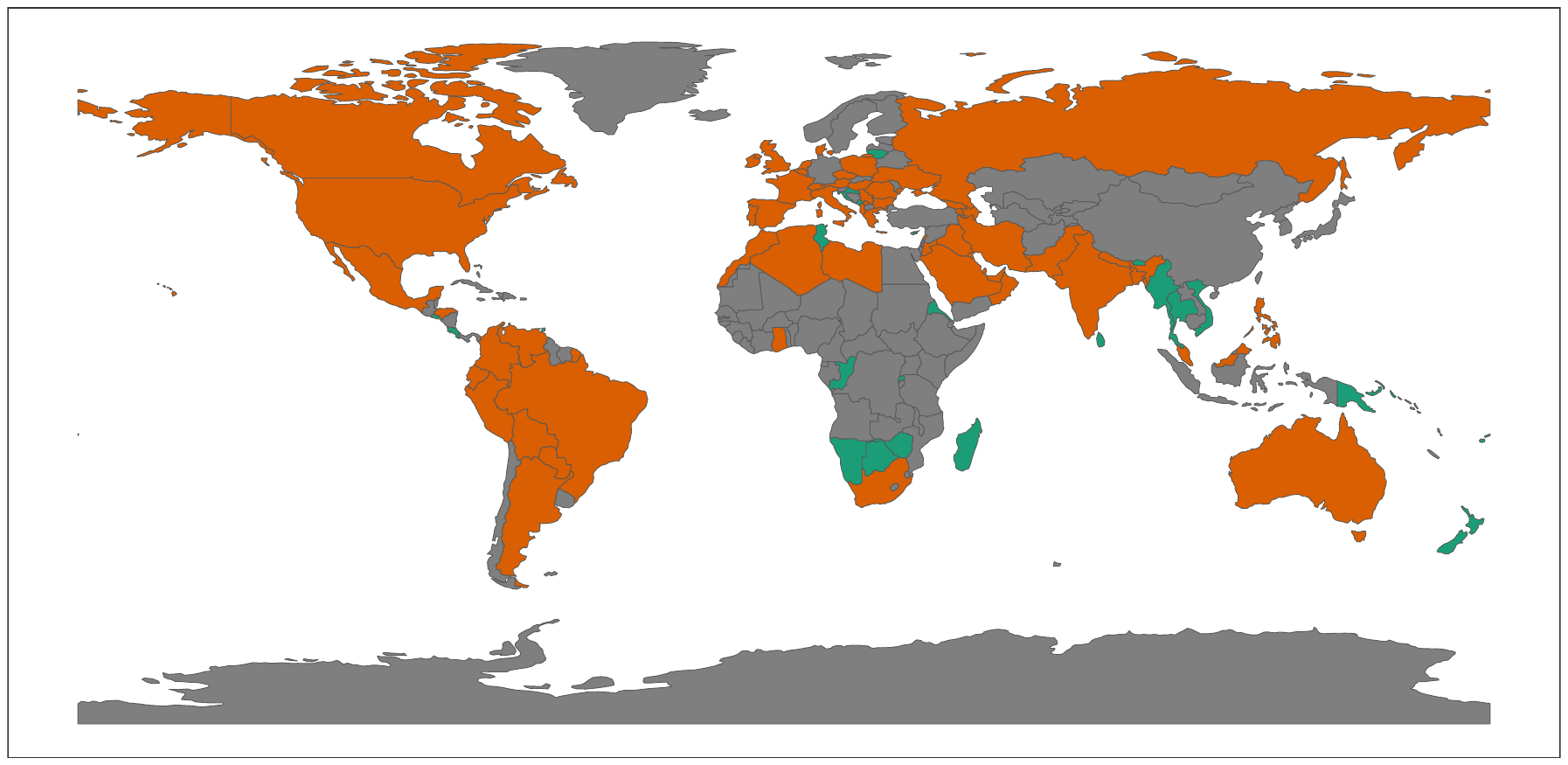

**Fig. S2.** Countries colored by whether their first lockdowns successfully contained COVID-19 initially.

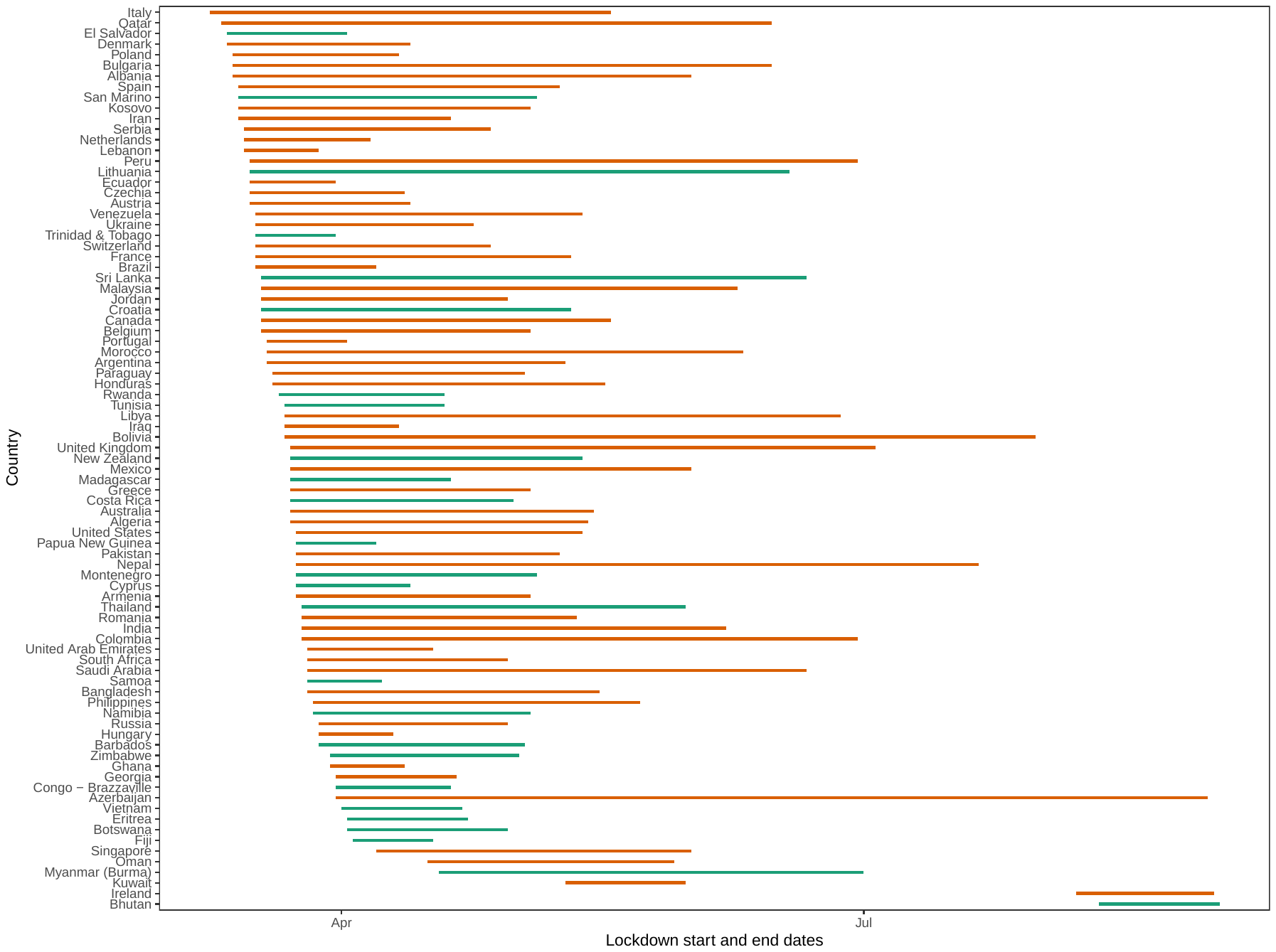

**Fig. S3.** Durations and start and end dates of lockdowns in various countries.

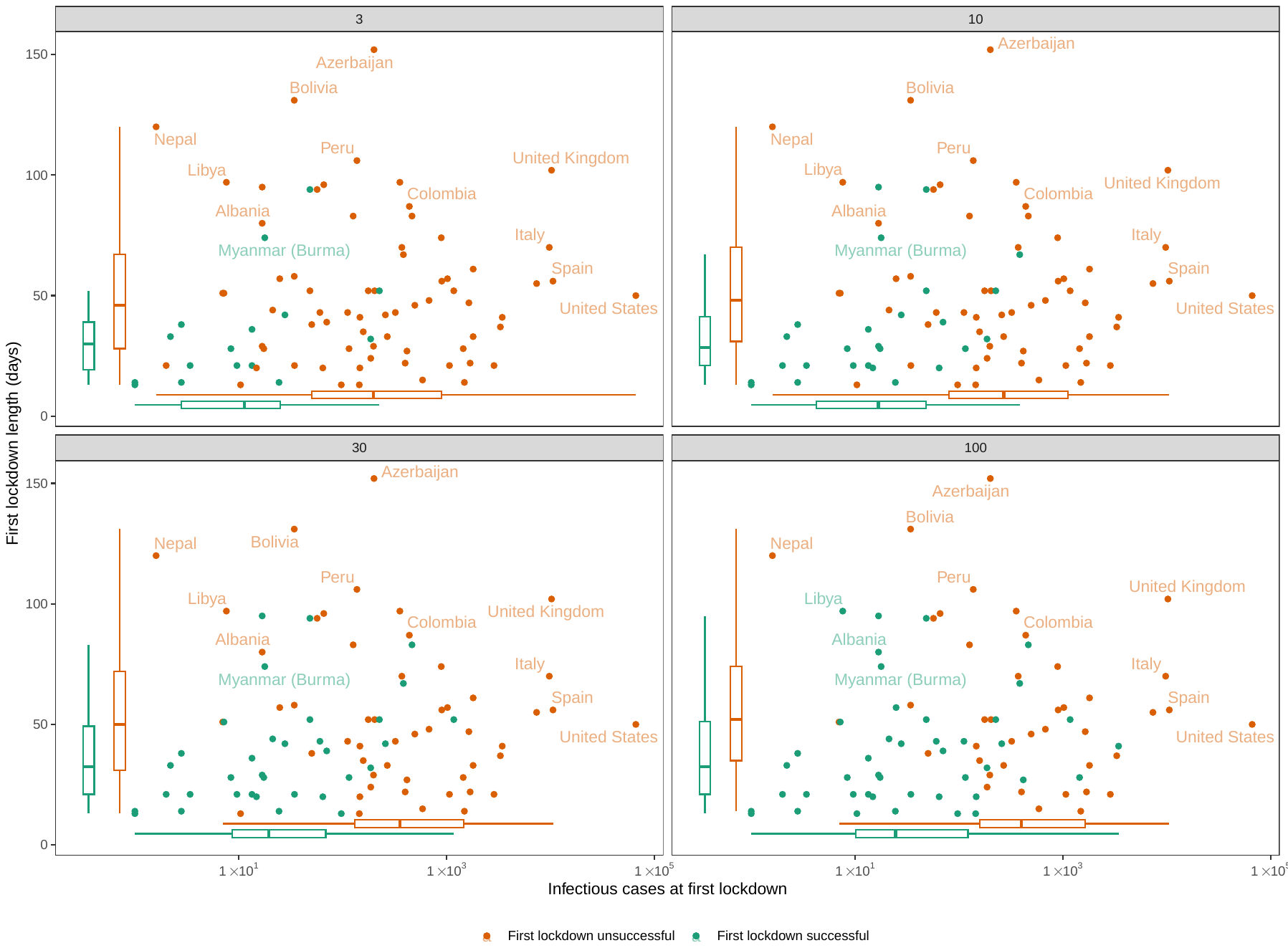

**Fig. S4.** Earliness of lockdown (x-axis) versus lockdown length in days (y-axis) and lockdown success (first lockdown unsuccessful (orange) and first lockdown successful (teal)), analogous to fig. S1, for 4 different thresholds of lockdown success (thresholds shown in gray labels). A lockdown is successful if the average number of daily cases following lockdown is less than the threshold for 7 days.

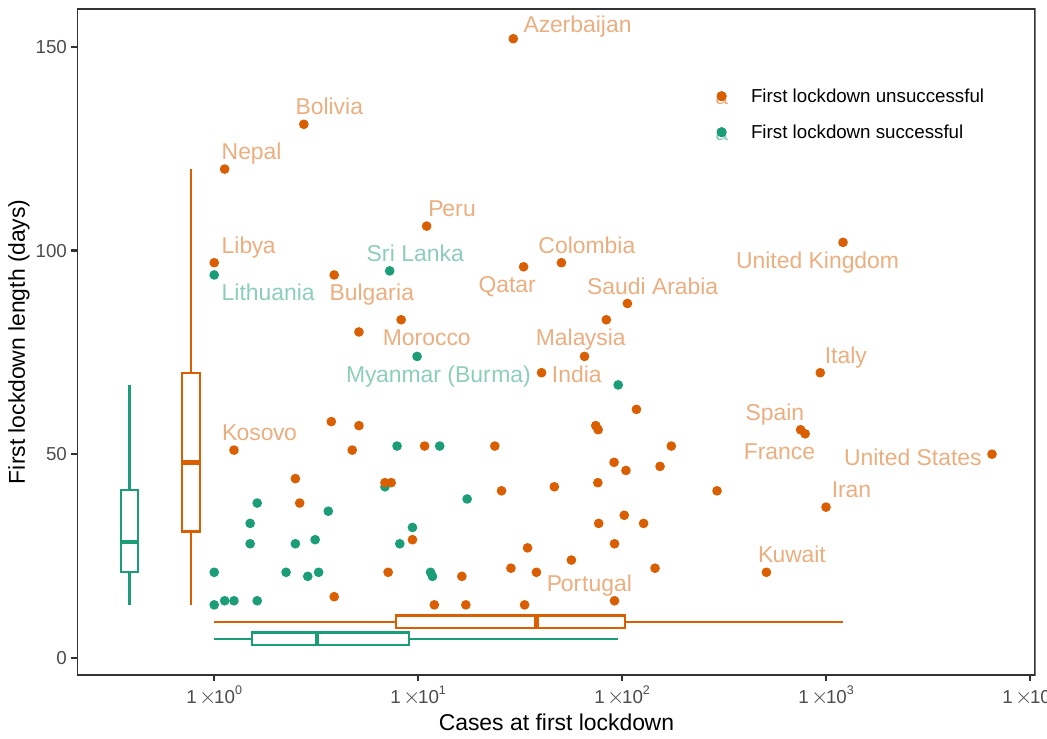

**Fig. S5.** Earliness of lockdown (x-axis) versus lockdown length in days (y-axis) and lockdown success (first lockdown unsuccessful (orange) and first lockdown successful (teal)), analogous to fig. S1, except that earliness of lockdown is measured here in terms of all cases (rather than infectious cases) at lockdown.

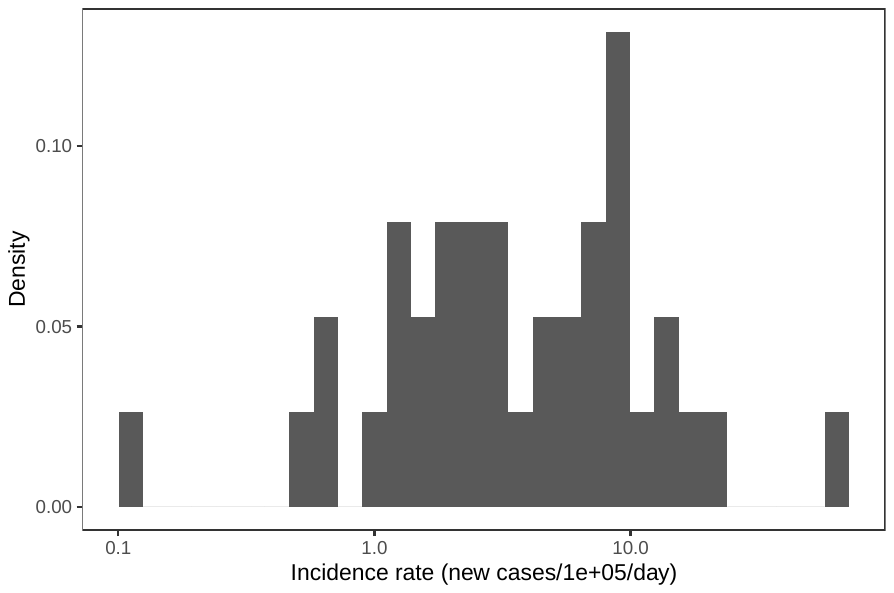

**Fig. S6.** Distribution of wastewater sensitivity estimated from reported COVID-19 incidences and wastewater sample data from 47 sampling locations from^53^.

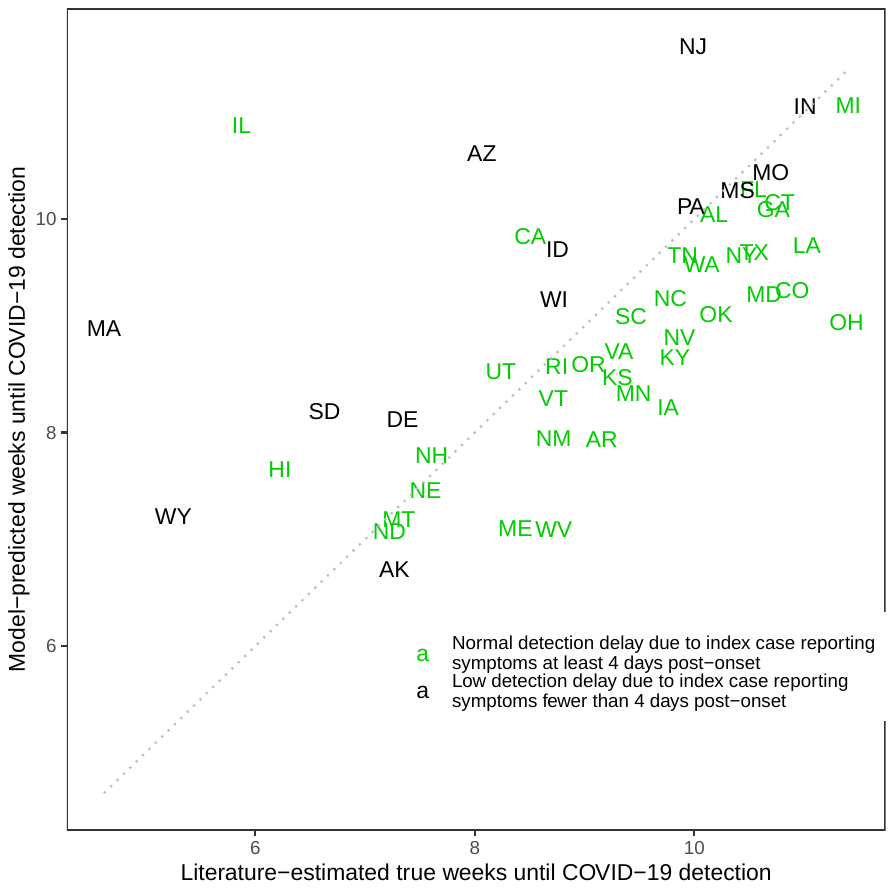

**Fig. S7.** Actual versus predicted COVID-19 weeks required to trigger each US state’s detection of its first 2020 COVID-19 outbreak (by monitoring symptomatic travel-associated cases).

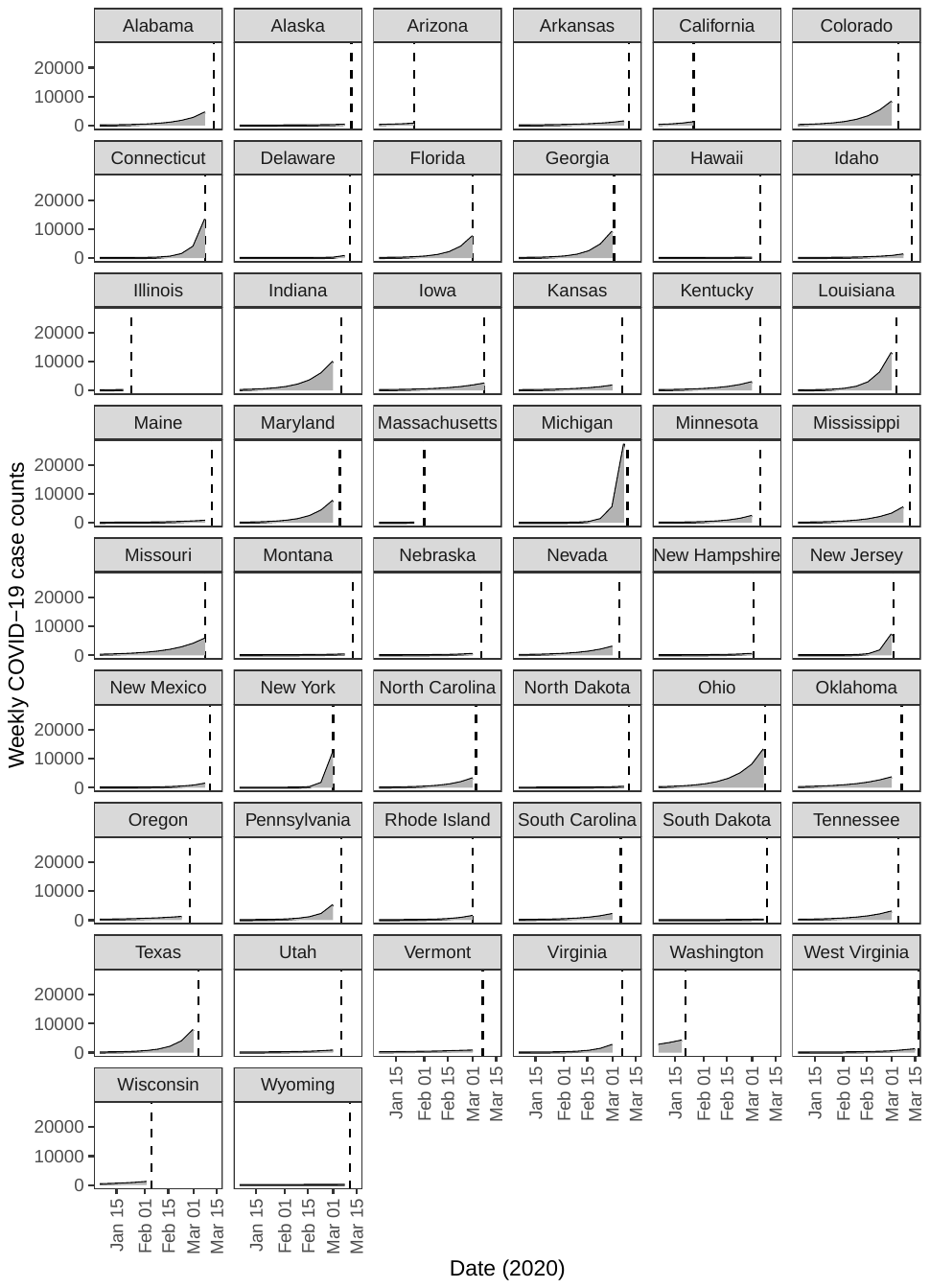

**Fig. S8.** COVID-19 cases leading up to COVID-19 detection in 50 US states, which are used as the x-axis in fig. S7. Y-values here are literature estimates of total (tested plus untested) cases^54^. We extrapolated these cases based on exponential fit back to January 1, 2020. Dashed lines mark the date of the first detected case in the state, and the shaded areas under the curve denote the cumulative number of cases until detection.

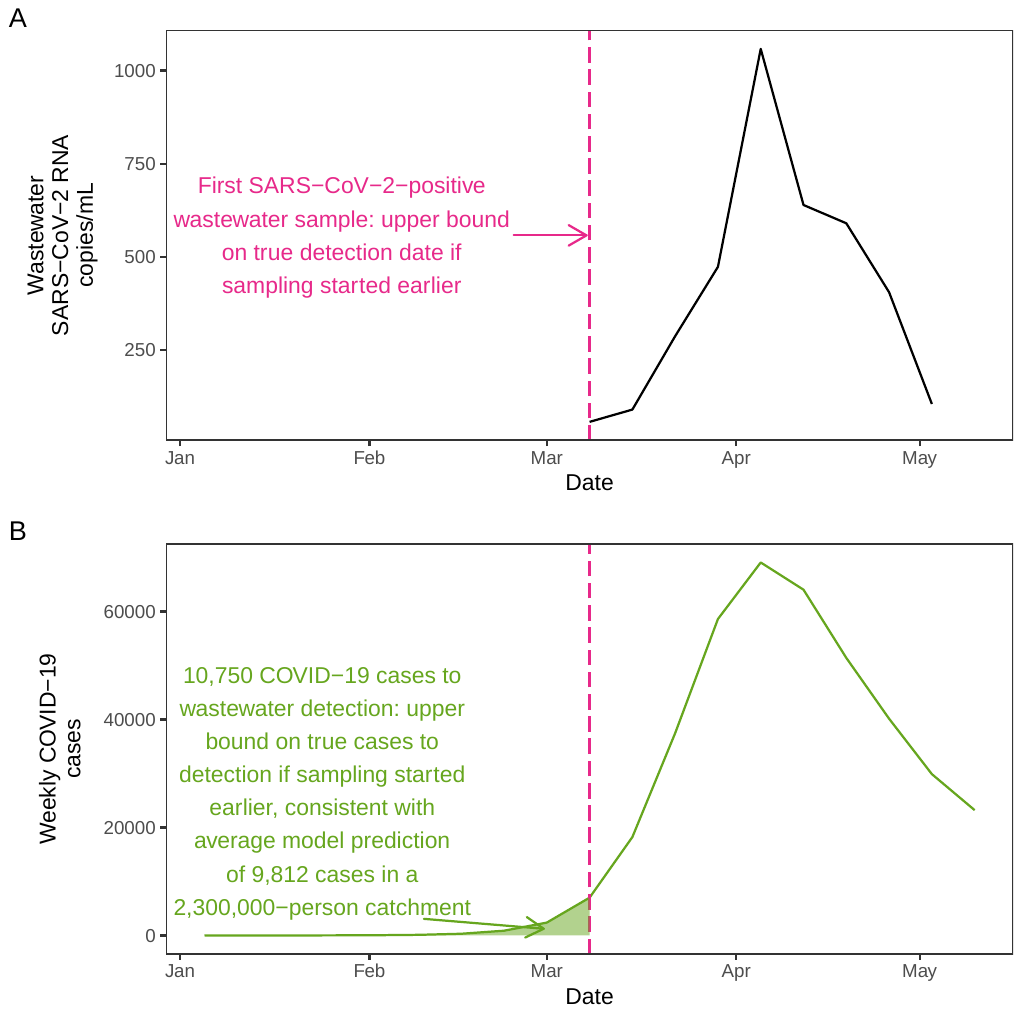

**Fig. S9.** Number of COVID-19 cases in Massachusetts before wastewater detection in 2020. (**A**) 7-day averages of SARS-CoV-2 RNA copies/mL in wastewater from the Deer Island Treatment Plant (combined Southern and Northern plants), the Massachusetts Water Resources Authority (MWRA) plant treating wastewater from 3.1 million people in the Boston metropolitan area in the United States (based on^57^). (**B**) Weekly non-cumulative COVID-19 case counts in the MWRA-covered communities (based on literature estimates of total tested and untested cases in^54^).

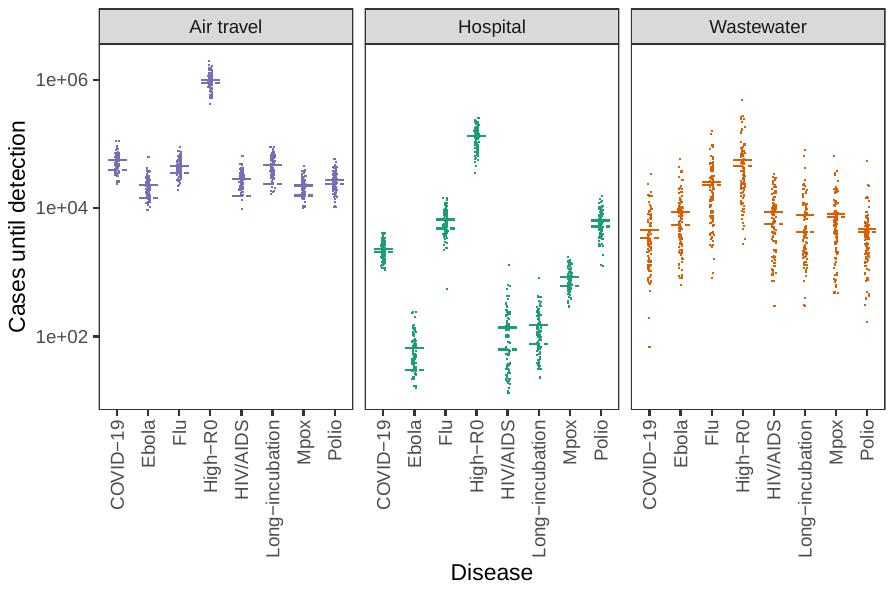

**Fig. S10.** Comparison of simulation model of cases until detection versus mathematical approximation (hospital (teal), wastewater (orange) and air travel (purple)) in a 650,000-person catchment. Solid lines are the means of simulated case counts; dashed lines are the approximated means based on the derived formula for cases at detection.

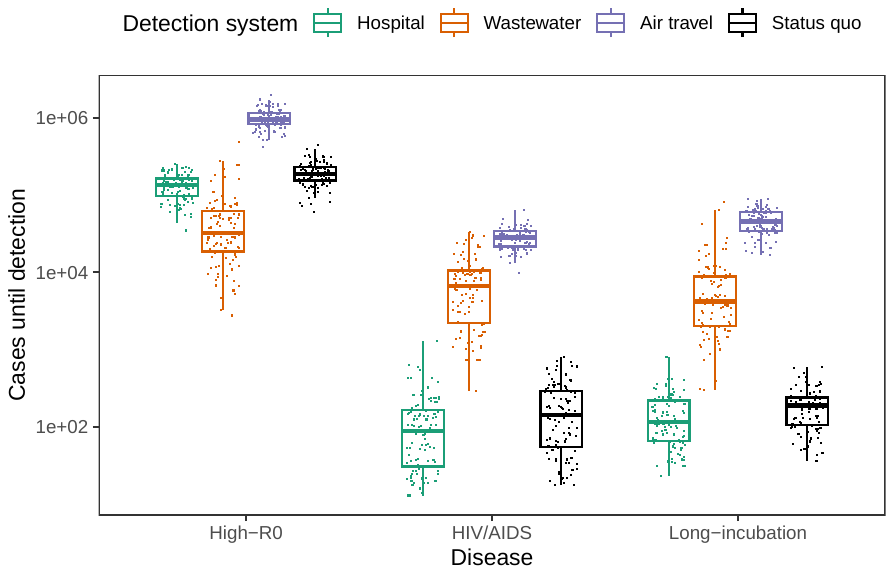

**Fig. S11.** Earliness of detection for detection systems in cases for additional infectious diseases in a 650,000-person catchment, akin to Fig. 2A (hospital (teal), wastewater (orange), air travel (purple) and status quo (black)).

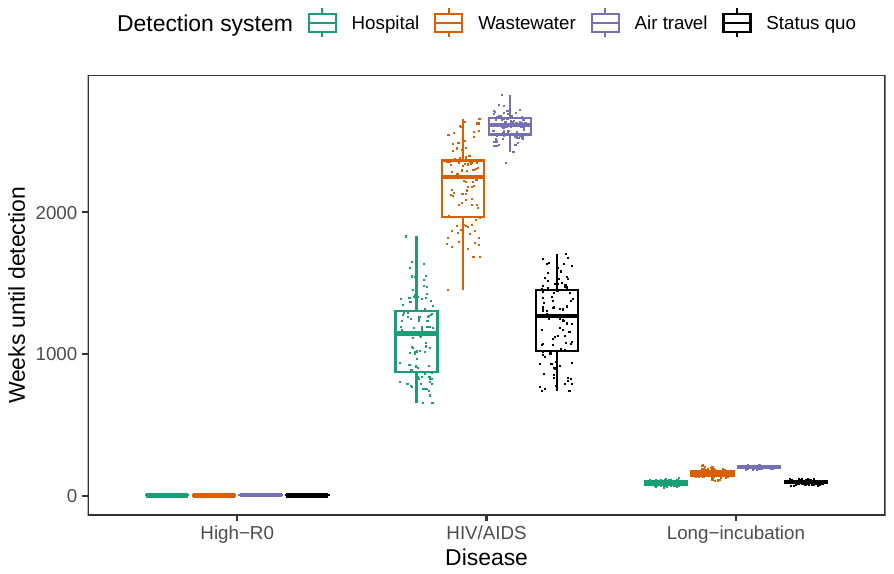

**Fig. S12.** Earliness of detection for detection systems in weeks for additional infectious diseases in a 650,000-person catchment, akin to Fig. 2A (hospital (teal), wastewater (orange), air travel (purple) and status quo (black)).

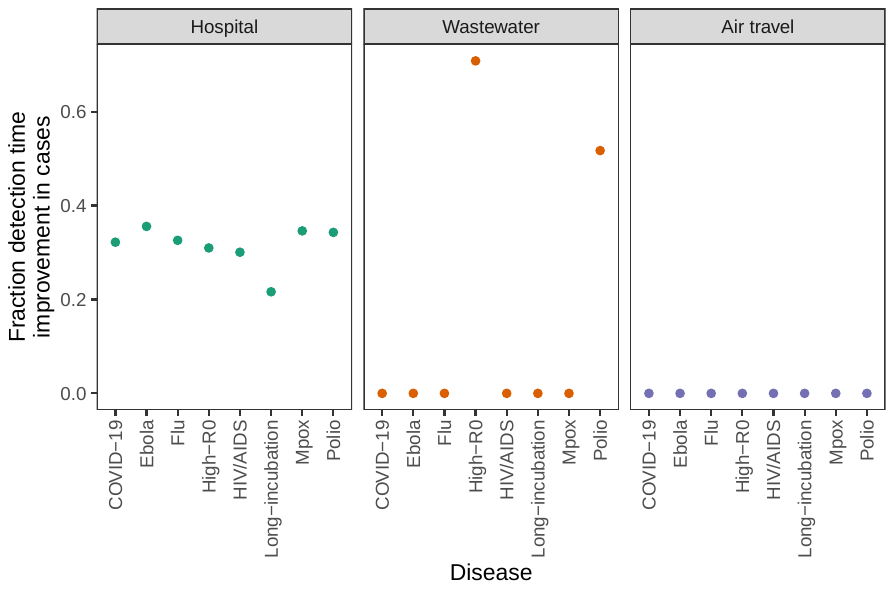

**Fig. S13.** Percent improvement in detection times in cases of the proposed systems over status quo detection for multiple outbreaks corresponding to Fig. 2A (air travel (purple), hospital (teal) and wastewater (orange)). Point shows percent improvement of mean system detection time over mean status quo detection time.

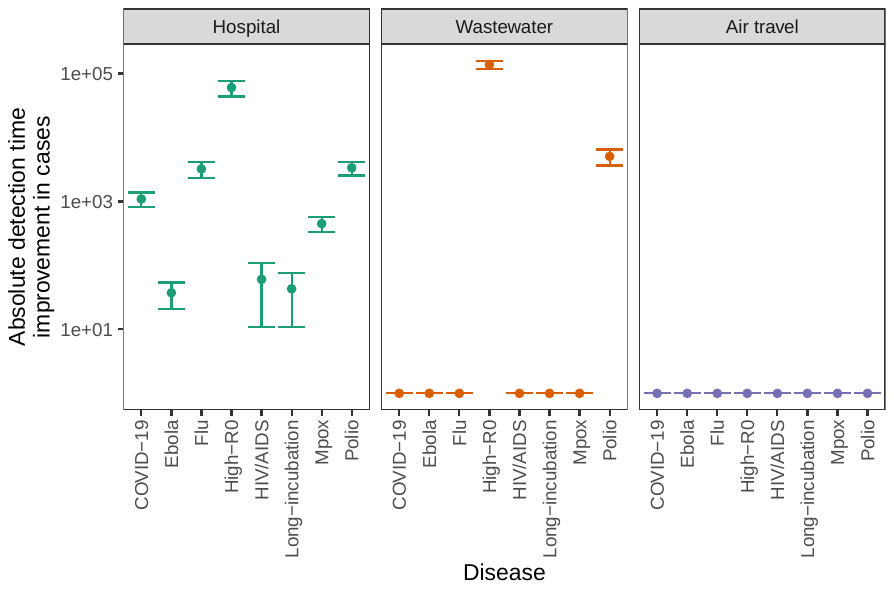

**Fig. S14.** Absolute improvement in detection times in cases of the proposed systems over status quo detection for multiple outbreaks corresponding to Fig. 2A (air travel (purple), hospital (teal) and wastewater (orange)). Bar shows 95% confidence interval from 2-sample 2-sided t-test between status quo and system detection times, and point shows interval midpoint or difference of means.

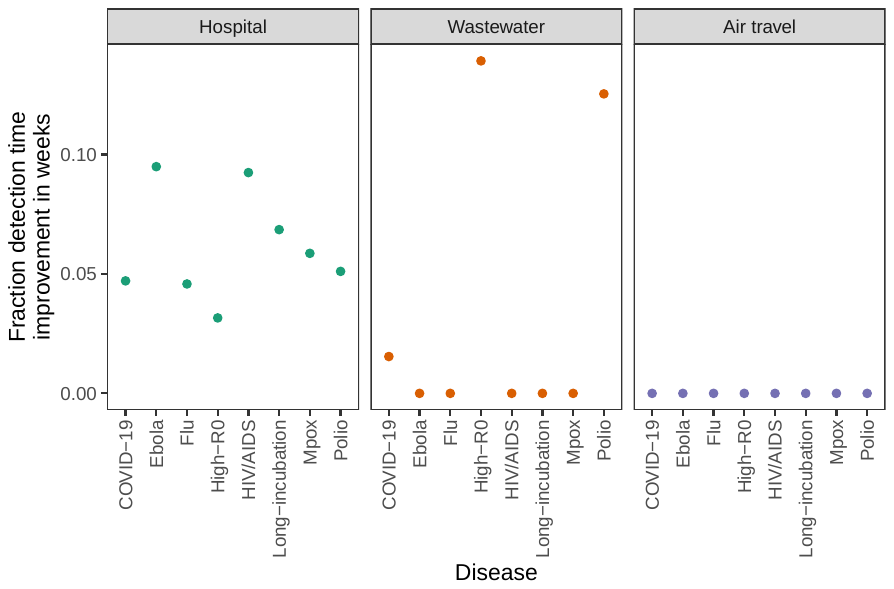

**Fig. S15.** Percent improvement in detection times in weeks of the proposed systems over status quo detection for multiple outbreaks corresponding to Fig. 2A (air travel (purple), hospital (teal) and wastewater (orange)). Point shows percent improvement of mean system detection time over mean status quo detection time.

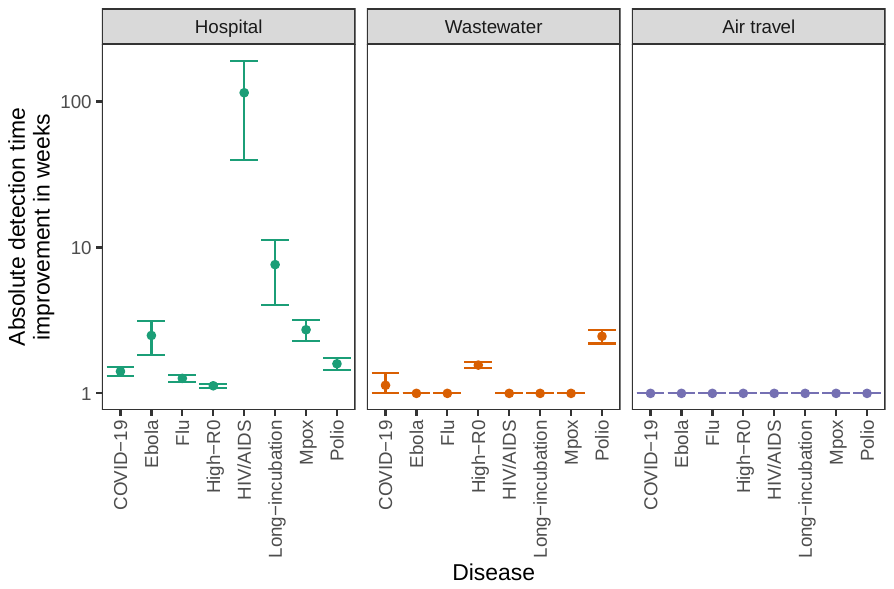

**Fig. S16.** Absolute improvement in detection times in weeks of the proposed systems over status quo detection for multiple outbreaks corresponding to Fig. 2A (air travel (purple), hospital (teal) and wastewater (orange)). Bar shows 95% confidence interval from 2-sample 2-sided t-test between status quo and system detection times, and point shows interval midpoint or difference of means.

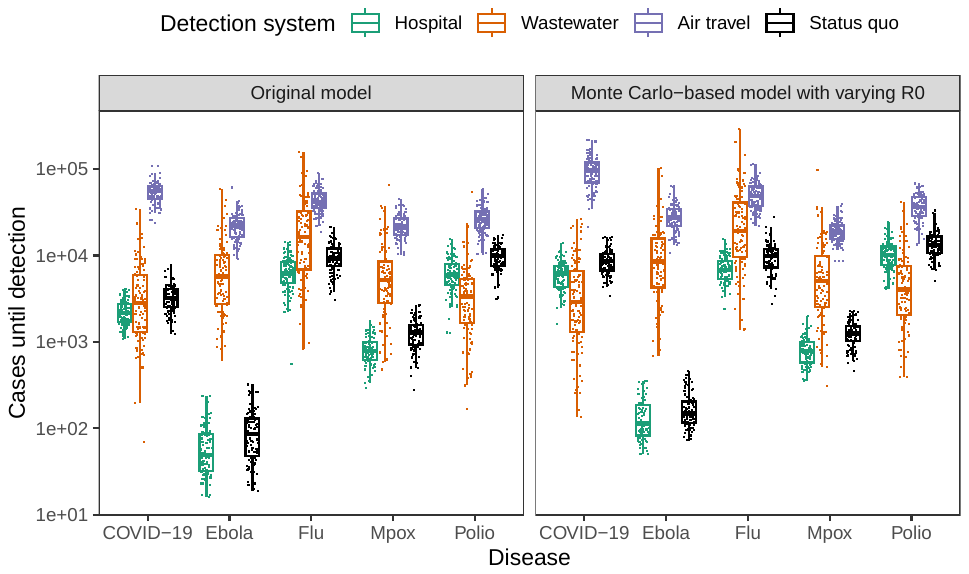

**Fig. S17.** Detection times estimated by main model versus Monte Carlo-based model with reproduction number^55^. The left panel is our original Fig. 2A using the original model; the right panel shows the same detection times from the more complex model.

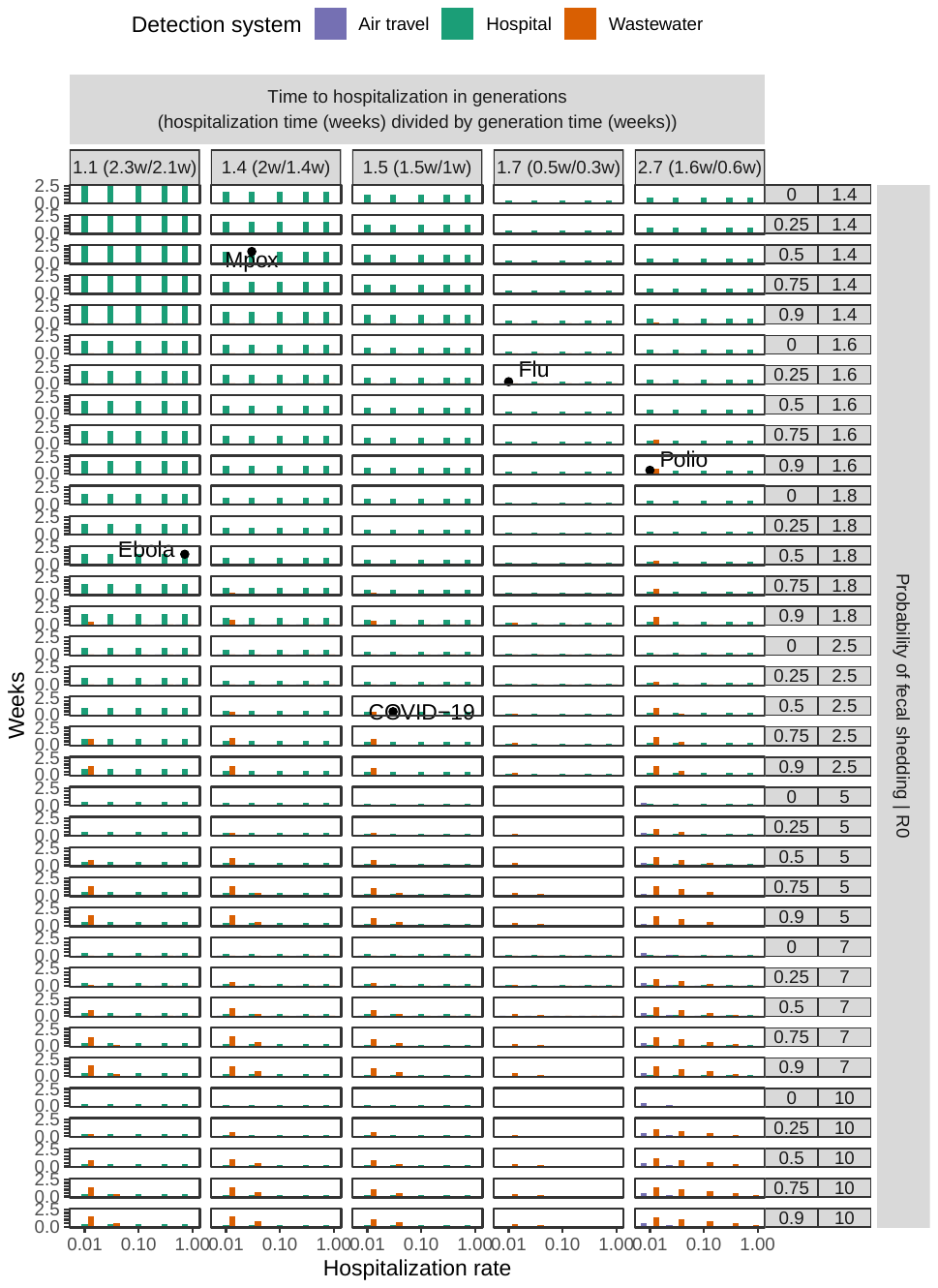

**Fig. S18.** Average weeks gained over status quo detection by the proposed detection systems across the epidemiological space of possible diseases. This is similar to Fig. 3.

**Table S1.** **Dates of first COVID-19 lockdowns implemented by 85 countries in 2020.** Dates are sourced from media reports.

| Country/Region | First lockdown start date | First lockdown end date | First lockdown length (days) |
| --- | --- | --- | --- |
| Albania | 2020-03-13 | 2020-06-01 | 80 |
| Algeria | 2020-03-23 | 2020-05-14 | 52 |
| Argentina | 2020-03-19 | 2020-05-10 | 52 |
| Armenia | 2020-03-24 | 2020-05-04 | 41 |
| Australia | 2020-03-23 | 2020-05-15 | 52 |
| Austria | 2020-03-16 | 2020-04-13 | 28 |
| Azerbaijan | 2020-03-31 | 2020-08-30 | 152 |
| Bangladesh | 2020-03-26 | 2020-05-16 | 51 |
| Barbados | 2020-03-28 | 2020-05-03 | 36 |
| Belgium | 2020-03-18 | 2020-05-04 | 47 |
| Bhutan | 2020-08-11 | 2020-09-01 | 21 |
| Bolivia | 2020-03-22 | 2020-07-31 | 131 |
| Botswana | 2020-04-02 | 2020-04-30 | 28 |
| Brazil | 2020-03-17 | 2020-04-07 | 21 |
| Bulgaria | 2020-03-13 | 2020-06-15 | 94 |
| Canada | 2020-03-18 | 2020-05-18 | 61 |
| Colombia | 2020-03-25 | 2020-06-30 | 97 |
| Congo - Brazzaville | 2020-03-31 | 2020-04-20 | 20 |
| Costa Rica | 2020-03-23 | 2020-05-01 | 39 |
| Croatia | 2020-03-18 | 2020-05-11 | 32 |
| Cyprus | 2020-03-24 | 2020-04-13 | 20 |
| Czechia | 2020-03-16 | 2020-04-12 | 27 |
| Denmark | 2020-03-12 | 2020-04-13 | 33 |
| Ecuador | 2020-03-16 | 2020-03-31 | 15 |
| El Salvador | 2020-03-12 | 2020-04-02 | 21 |
| Eritrea | 2020-04-02 | 2020-04-23 | 21 |
| Fiji | 2020-04-03 | 2020-04-17 | 14 |
| France | 2020-03-17 | 2020-05-11 | 55 |
| Georgia | 2020-03-31 | 2020-04-21 | 21 |
| Ghana | 2020-03-30 | 2020-04-12 | 13 |
| Greece | 2020-03-23 | 2020-05-04 | 42 |
| Honduras | 2020-03-20 | 2020-05-17 | 58 |
| Hungary | 2020-03-28 | 2020-04-10 | 13 |
| India | 2020-03-25 | 2020-06-07 | 74 |
| Iran | 2020-03-14 | 2020-04-20 | 37 |
| Iraq | 2020-03-22 | 2020-04-11 | 20 |
| Ireland | 2020-08-07 | 2020-08-31 | 24 |
| Italy | 2020-03-09 | 2020-05-18 | 70 |
| Jordan | 2020-03-18 | 2020-04-30 | 43 |
| Kosovo | 2020-03-14 | 2020-05-04 | 51 |
| Kuwait | 2020-05-10 | 2020-05-31 | 21 |
| Lebanon | 2020-03-15 | 2020-03-28 | 13 |
| Libya | 2020-03-22 | 2020-06-27 | 97 |
| Lithuania | 2020-03-16 | 2020-06-18 | 94 |
| Madagascar | 2020-03-23 | 2020-04-20 | 28 |
| Malaysia | 2020-03-18 | 2020-06-09 | 83 |
| Mexico | 2020-03-23 | 2020-06-01 | 70 |
| Montenegro | 2020-03-24 | 2020-05-05 | 42 |
| Morocco | 2020-03-19 | 2020-06-10 | 83 |
| Myanmar (Burma) | 2020-04-18 | 2020-07-01 | 74 |
| Namibia | 2020-03-27 | 2020-05-04 | 38 |
| Nepal | 2020-03-24 | 2020-07-21 | 120 |
| Netherlands | 2020-03-15 | 2020-04-06 | 22 |
| New Zealand | 2020-03-23 | 2020-05-13 | 52 |
| Oman | 2020-04-16 | 2020-05-29 | 43 |
| Pakistan | 2020-03-24 | 2020-05-09 | 46 |
| Papua New Guinea | 2020-03-24 | 2020-04-07 | 14 |
| Paraguay | 2020-03-20 | 2020-05-03 | 44 |
| Peru | 2020-03-16 | 2020-06-30 | 106 |
| Philippines | 2020-03-27 | 2020-05-23 | 57 |
| Poland | 2020-03-13 | 2020-04-11 | 29 |
| Portugal | 2020-03-19 | 2020-04-02 | 14 |
| Qatar | 2020-03-11 | 2020-06-15 | 96 |
| Romania | 2020-03-25 | 2020-05-12 | 48 |
| Russia | 2020-03-28 | 2020-04-30 | 33 |
| Rwanda | 2020-03-21 | 2020-04-19 | 29 |
| Samoa | 2020-03-26 | 2020-04-08 | 13 |
| San Marino | 2020-03-14 | 2020-05-05 | 52 |
| Saudi Arabia | 2020-03-26 | 2020-06-21 | 87 |
| Serbia | 2020-03-15 | 2020-04-27 | 43 |
| Singapore | 2020-04-07 | 2020-06-01 | 56 |
| South Africa | 2020-03-26 | 2020-04-30 | 35 |
| Spain | 2020-03-14 | 2020-05-09 | 56 |
| Sri Lanka | 2020-03-18 | 2020-06-21 | 95 |
| Switzerland | 2020-03-17 | 2020-04-27 | 41 |
| Thailand | 2020-03-25 | 2020-05-31 | 67 |
| Trinidad & Tobago | 2020-03-17 | 2020-03-31 | 14 |
| Tunisia | 2020-03-22 | 2020-04-19 | 28 |
| Ukraine | 2020-03-17 | 2020-04-24 | 38 |
| United Arab Emirates | 2020-03-26 | 2020-04-17 | 22 |
| United Kingdom | 2020-03-23 | 2020-07-03 | 102 |
| United States | 2020-03-24 | 2020-05-13 | 50 |
| Venezuela | 2020-03-17 | 2020-05-13 | 57 |
| Vietnam | 2020-04-01 | 2020-04-22 | 21 |
| Zimbabwe | 2020-03-30 | 2020-05-02 | 33 |

**Table S2.** **Threshold, delay, and probability for 3 proposed early detection systems.**

| Detection system | Detection probability | Detection threshold | Detection delay |
| --- | --- | --- | --- |
| Hospital ^1^ | outbreak hospitalization rate | 10 | (outbreak time to hospitalization (weeks) + logistical detection delay)/outbreak serial interval (weeks) |
| Wastewater ^2^ | fraction of people connected to central sewage * outbreak probability of fecal shedding | adjusted draw from^53^ (median 2.5e-5) * community catchment population (non-cumulative) | (outbreak time to fecal shedding (weeks) + logistical detection delay)/outbreak serial interval (weeks) |
| Air travel ^3^ | weekly probability of international travel * symptomatic rate | 10 | (outbreak serial interval (weeks) + logistical detection delay)/outbreak serial interval (weeks) |

^1^ Detection threshold: The government or hospital implementing the system chooses the detection threshold they consider to be sufficient. For COVID-19, Wuhan hospitals were willing to report the “extraordinary” situation to local health authorities after seven known cases^69^. During the 2002-2004 SARS-CoV-1 outbreak, hospital officials became alarmed after one patient and eight doctors and nurses became sick^71^. Thus we choose a detection threshold of ten.

^2^ See Materials and methods for details on setting the detection threshold. Detection delay:^63,76^. Detection probability: Fecal shedding tends to constitute more of the human pathogen nucleic acid in wastewater than urine, saliva, or other specimens, due to higher rates of shedding and higher pathogen loads in feces^63,77^. The fraction of people connected to central sewage in Wuhan is estimated at 80% based on a 2016 Asian Development Bank appraisal stating that Wuhan aimed to treat this fraction of wastewater in 2010^78^; this fraction is similar to the fraction of US households connected to public sewers (83%)^64^.

^3^ Detection threshold: reasoning is similar to hospital monitoring reasoning for detection threshold.

**Table S3.** **Epidemiological parameters of outbreaks studied.**

| Outbreak | Hospitalization rate | R0 | Serial interval (weeks) | Time to hospitalization (weeks) | Probability of fecal shedding | Dispersion |
| --- | --- | --- | --- | --- | --- | --- |
| COVID-19 ^1^ | 0.03 | 2.5 | 1.0 | 1.5 | 0.50 | 0.7 |
| Mpox (2022) ^2^ | 0.03 | 1.4 | 1.4 | 2.0 | 0.50 | 0.1 |
| Polio (2013-2014) ^3^ | 0.01 | 1.6 | 0.6 | 1.6 | 0.90 | 0.1 |
| Ebola (2013-2016) ^4^ | 0.72 | 1.8 | 2.1 | 2.3 | 0.50 | 0.1 |
| Flu (2009 pandemic) ^5^ | 0.01 | 1.6 | 0.3 | 0.5 | 0.25 | 0.1 |
| High-R0 (hypothetical) | 0.03 | 20.0 | 1.0 | 1.5 | 0.50 | 0.1 |
| HIV/AIDS (1980s-) ^6^ | 1.00 | 2.5 | 234.0 | 468.0 | 0.37 | 0.1 |
| Long-incubation (hypothetical) ^7^ | 0.50 | 3.0 | 20.8 | 25.0 | 0.50 | 0.1 |

^147,63,79,80^.

^248,81^. Due to the lack of *infection* hospitalization rates at this time, we infer the infection hospitalization rate to be 0.03 by halving the estimated *case* hospitalization rates of 0.06-0.07 for the 2022 mpox outbreak^82,83^. We choose half because a majority of mpox infections are symptomatic^84^ and some fraction of those will seek medical care and get tested. Time to hospitalization is estimated by adding the incubation period of 7 days to the median time from symptom onset to hospitalization (7 days)^85^. We and others are unable to find estimates of mpox fecal shedding rates^86^, but it has been detectable in wastewater during the 2022 mpox outbreak^87^, so we assign a value of 0.5, in line with SARS-CoV-2 and flu, but on the higher end because mpox causes symptoms more broadly than in just the respiratory system.

^3^ Due to lack of data and estimates of R0 for polio in 2022, we use an R0 of 1.6 from the Israel 2013-2014 wild poliovirus type 1 outbreak^60^ to represent a polio outbreak in a population with sanitation systems and high levels of vaccination coverage^88,89^. Hospitalization rate is inferred from the fact that less than or near 1% of polio infections result in flaccid paralysis^90^. Serial interval is estimated as the latent period plus one half of the infectious period^91^: in the Israel outbreak, this was estimated as $1/\sigma+1/2*1/\gamma=4+1/2*1/0.93\approx4.5$ days (Table 2 in^60^). Hospitalization time is inferred from the several-day period of minor illness, symptom-free period of 1-3 days, and then onset of paralysis within 2-3 days^90^. Probability of fecal shedding was inferred from literature estimates in enteroviruses^92^ and in vaccinated children^93^.

^494,95^. The time to hospitalization is estimated as the sum of the incubation period (9-12 days^96^) and the time from symptom onset to hospital admission (5.7 days^97^). We and others are unable to find precise estimates of Ebola fecal shedding rates, but Ebola has commonly been detected in stool when measured^77^, so we assign a value of 0.5, in line with SARS-CoV-2 and flu, but on the higher end because Ebola causes symptoms more broadly than in just the respiratory system.

^598,99^. The hospitalization rate was estimated by multiplying the symptomatic hospitalization rate of 0.0144 (the proportion of symptomatic cases requiring hospitalization)^100^ by the symptomatic rate of 0.8 (the proportion of all cases who were symptomatic)^101^. The hospitalization time was estimated as the sum of the incubation period (1.4 days^102^) and the time from symptom onset to hospital admission (2 days^103^).

^6104–107^. Probability of fecal shedding is calculated using estimates that 60% of HIV-positive patients show gastrointestinal symptoms^108^ and 5/9 and 1/10 of HIV-positive patients showing and not showing gastrointestinal symptoms, respectively, test positive in fecal samples for HIV nucleic acid^109^.

^7^ These parameters are very loosely inspired by the parameters for long-incubation diseases like tuberculosis (assuming cases are untreated)^110–112^. Time to active disease is used as a proxy for time to hospitalization. The serial interval is estimated by taking estimates from the antibiotic era and subtracting 12 months to account for 12 months of antibiotics treatment, and this is consistent with the observed pre-antibiotic era incubation period of at least 1-1.5 months (assuming transmission starts approximately when symptoms appear), because the serial interval is the latent period plus half the infectious period. Reproductive number is selected from the higher end of^112^ because most of the studies in that review are from the antibiotic era.

**Table S4.** **Date of first reported COVID-19 case in each of 50 US states.** Dates are sourced from media reports and state public health agency press releases. An index case is considered to be caught unusually early if caught earlier than 4 days after symptom onset.

| Location | Postal code | First case date | Index case caught unusually early |
| --- | --- | --- | --- |
| Washington ^1^ | WA | 2020-01-21 | FALSE |
| Illinois ^2^ | IL | 2020-01-24 | FALSE |
| Arizona ^3^ | AZ | 2020-01-26 | TRUE |
| California ^4^ | CA | 2020-01-26 | FALSE |
| Massachusetts ^5^ | MA | 2020-02-01 | TRUE |
| Wisconsin ^6^ | WI | 2020-02-05 | TRUE |
| Oregon ^7^ | OR | 2020-02-28 | FALSE |
| New York ^8^ | NY | 2020-03-01 | FALSE |
| Florida ^9^ | FL | 2020-03-01 | FALSE |
| Rhode Island ^10^ | RI | 2020-03-01 | FALSE |
| Georgia ^11^ | GA | 2020-03-02 | FALSE |
| New Hampshire ^12^ | NH | 2020-03-02 | FALSE |
| New Jersey ^13^ | NJ | 2020-03-02 | TRUE |
| North Carolina ^14^ | NC | 2020-03-03 | FALSE |
| Louisiana ^15^ | LA | 2020-03-04 | FALSE |
| Texas ^16^ | TX | 2020-03-04 | FALSE |
| Maryland ^17^ | MD | 2020-03-05 | FALSE |
| Colorado ^18^ | CO | 2020-03-05 | FALSE |
| Tennessee ^19^ | TN | 2020-03-05 | FALSE |
| Nevada ^20^ | NV | 2020-03-05 | FALSE |
| Hawaii ^21^ | HI | 2020-03-06 | FALSE |
| Minnesota ^22^ | MN | 2020-03-06 | FALSE |
| Utah ^23^ | UT | 2020-03-06 | FALSE |
| Nebraska ^24^ | NE | 2020-03-06 | FALSE |
| Indiana ^25^ | IN | 2020-03-06 | TRUE |
| Pennsylvania ^26^ | PA | 2020-03-06 | TRUE |
| Kentucky ^27^ | KY | 2020-03-06 | FALSE |
| South Carolina ^28^ | SC | 2020-03-06 | FALSE |
| Virginia ^29^ | VA | 2020-03-07 | FALSE |
| Oklahoma ^30^ | OK | 2020-03-07 | FALSE |
| Kansas ^31^ | KS | 2020-03-07 | FALSE |
| Vermont ^32^ | VT | 2020-03-07 | FALSE |
| Iowa ^33^ | IA | 2020-03-08 | FALSE |
| Missouri ^34^ | MO | 2020-03-08 | TRUE |
| Connecticut ^35^ | CT | 2020-03-08 | FALSE |
| Ohio ^36^ | OH | 2020-03-09 | FALSE |
| Michigan ^37^ | MI | 2020-03-10 | FALSE |
| South Dakota ^38^ | SD | 2020-03-10 | TRUE |
| New Mexico ^39^ | NM | 2020-03-11 | FALSE |
| North Dakota ^40^ | ND | 2020-03-11 | FALSE |
| Arkansas ^41^ | AR | 2020-03-11 | FALSE |
| Delaware ^42^ | DE | 2020-03-11 | TRUE |
| Wyoming ^43^ | WY | 2020-03-11 | TRUE |
| Maine ^44^ | ME | 2020-03-12 | FALSE |
| Alaska ^45^ | AK | 2020-03-12 | TRUE |
| Mississippi ^46^ | MS | 2020-03-12 | TRUE |
| Idaho ^47^ | ID | 2020-03-13 | TRUE |
| Alabama ^48^ | AL | 2020-03-13 | FALSE |
| Montana ^49^ | MT | 2020-03-13 | FALSE |
| West Virginia ^50^ | WV | 2020-03-17 | FALSE |

^1^ <https://www.seattletimes.com/seattle-news/health/case-of-wuhan-coronavirus-detected-in-washington-state-first-in-united-states/>

^2^ <https://www.ncbi.nlm.nih.gov/pmc/articles/PMC7158585/> <https://www.cbsnews.com/chicago/news/first-case-of-coronavirus-confirmed-in-chicago/>

^3^ <https://www.thedailybeast.com/5th-us-case-of-coronavirus-confirmed-in-arizona>; <https://www.azcentral.com/story/news/local/arizona-breaking/2020/01/26/first-case-coronavirus-reaches-arizona-fifth-person-infected/4582588002/>; <https://www.azcentral.com/story/news/local/arizona-health/2020/02/21/arizona-only-new-coronavirus-patient-out-isolation/4836396002/>

^4^ <https://www.ocregister.com/2020/01/26/coronavirus-patients-confirmed-in-los-angeles-and-orange-counties/>

^5^ <https://www.bostonherald.com/2020/02/01/first-case-of-coronavirus-confirmed-in-massachusetts-dph/>

^6^ <https://www.wkow.com/coronavirus/first-case-of-coronavirus-in-wisconsin-confirmed/article_1776a5a8-6356-5da1-ac36-80d672a44188.html>

^7^ <https://www.oregonlive.com/news/2020/02/coronavirus-appears-in-oregon.html> <https://pamplinmedia.com/lor/48-news/465562-377333-forest-hills-employee-recovering-from-coronavirus>

^8^ <https://www.wsj.com/articles/first-case-of-coronavirus-confirmed-in-new-york-state-11583111692> <https://edition.cnn.com/2020/03/02/us/new-york-coronavirus-first-case/index.html>

^9^ <https://www.wesh.com/article/coronavirus-cases-florida-public-health-emergency/31189479>

^10^ <https://www.providencejournal.com/story/special/2020/03/04/ris-first-case-of-coronavirus-confirmed-precautions-broaden/1573947007/> <https://www.pressherald.com/2020/03/01/first-virus-case-confirmed-in-rhode-island/>

^11^ <https://www.11alive.com/article/news/health/fulton-county-coronavirus-cases/85-9f552f0f-5d2f-4043-9b7c-738d914b8d1a> <https://www.wsbtv.com/news/local/first-cases-coronavirus-confirmed-georgia/4P22YK37OBF2ZIC5VY2YOX7KDE/>

^12^ <https://www.nhpr.org/health/2020-03-02/first-positive-test-results-for-coronavirus-identified-in-n-h>

^13^ <https://www.northjersey.com/story/news/bergen/2020/03/04/nj-gov-phil-murphy-bergen-county-nj-first-confirmed-case-coronavirus-covid-19/4958681002/> <https://www.fox5ny.com/news/new-jersey-announces-first-presumptive-case-of-covid-19-coronavirus> <https://www.northjersey.com/story/news/health/2020/03/10/coronavirus-new-jersey-timeline-events-covid-covid-19/4964918002/>

^14^ <https://www.ncdhhs.gov/news/press-releases/2020/03/03/north-carolina-identifies-first-case-covid-19> <https://abc11.com/coronavirus-nc-north-carolina-symptoms/5982249/>

^15^ <https://www.theadvertiser.com/story/news/2020/03/09/coronavirus-louisianas-first-patient-being-treated-pneumonia-hospital/5004688002/>

^16^ <https://www.texastribune.org/2020/03/04/texas-coronavirus-case-confirmed-fort-bend-county/>

^17^ <https://www.washingtonpost.com/world/middle_east/new-coronavirus-cluster-linked-to-nile-river-cruise-ship-popular-with-tourists/2020/03/06/33c79b24-5fae-11ea-ac50-18701e14e06d_story.html> <https://www.washingtonpost.com/local/md-politics/maryland-confirms-three-cases-of-coronavirus/2020/03/05/687def10-5f3d-11ea-b014-4fafa866bb81_story.html>

^18^ <https://www.colorado.gov/governor/news/updated-information-covid-19> <https://coloradosun.com/2020/03/31/el-paso-county-coronavirus-deaths/>

^19^ <https://www.tennessean.com/story/news/health/2020/03/05/coronavirus-tennessee-department-health-bill-lee/4962037002/>

^20^ <https://www.staradvertiser.com/2020/03/05/breaking-news/tourist-mecca-las-vegas-sees-nevadas-first-coronavirus-case/>

^21^ <https://www.staradvertiser.com/2020/03/07/hawaii-news/hawaiis-first-case-of-coronavirus-confirmed-gov-david-ige-announces/>

^22^ <https://www.kare11.com/article/news/health/coronavirus/coronavirus-in-minnesota/89-da7d7967-ff29-4d69-ae12-079d12fa5752>

^23^ <https://health.utah.gov/featured-news/utah-health-officials-announce-first-case-of-covid-19>

^24^ <https://nebraskapublicmedia.org/en/news/news-articles/nebraska-confirms-first-covid-19-case/>

^25^ <https://www.indystar.com/story/news/health/2020/03/06/coronavirus-indiana-cases/4973737002/> <https://www.wrtv.com/coronavirus/first-case-of-coronavirus-confirmed-in-indiana>

^26^ <https://www.governor.pa.gov/newsroom/wolf-administration-confirms-two-presumptive-positive-cases-of-covid-19/> <https://www.delcotimes.com/2022/03/09/delaware-county-commemorates-the-1800-lost-to-covid/>

^27^ <https://kentucky.gov/Pages/Activity-stream.aspx?n=GovernorBeshear&prId=77> <https://www.lpm.org/news/2021-03-05/a-whole-other-reality-inside-the-first-confirmed-case-of-covid-19-in-kentucky>

^28^ <https://www.thestate.com/news/state/south-carolina/article240838221.html>

^29^ <https://www.cnn.com/2020/03/07/politics/us-marine-coronavirus-virginia/index.html> <https://www.vdh.virginia.gov/blog/2020/03/08/virginia-department-of-health-confirms-second-presumptive-positive-case-of-coronavirus-disease-2019-covid-19-in-state/>

^30^ <https://www.oklahoman.com/story/news/nation-world/2020/03/07/first-case-of-coronavirus-in-oklahoma-confirmed/60377613007/>

^31^ <https://www.kcur.org/health/2020-03-07/the-first-case-of-coronavirus-in-kansas-is-confirmed-in-johnson-county#stream/0>

^32^ <https://www.sevendaysvt.com/OffMessage/archives/2020/03/07/vermont-health-department-reports-the-states-first-coronavirus-case> <https://vtdigger.org/2020/03/07/vermont-has-first-presumptive-case-of-coronavirus/> <https://www.vermontpublic.org/vpr-news/2020-03-08/health-officials-announce-first-coronavirus-case-in-vermont>

^33^ <https://apnews.com/article/virus-outbreak-science-iowa-kim-reynolds-ia-state-wire-4fed9d219a5c62f884f4eae081ddfa9b>

^34^ <https://www.youtube.com/watch?v=98jpfNOw87I> <https://governor.mo.gov/press-releases/archive/governor-parson-state-and-local-officials-confirm-first-case-covid-19-test-0>

^35^ <https://www.nytimes.com/2020/03/08/nyregion/coronavirus-connecticut.html>

^36^ <https://www.cincinnati.com/story/news/2020/03/09/coronavirus-covid-19-ohio-positive-case/5003009002/>

^37^ <https://www.wxyz.com/news/coronavirus/first-cases-of-coronavirus-confirmed-in-michigan-one-each-in-oakland-and-wayne-counties>

^38^ <https://thehill.com/policy/healthcare/486917-south-dakota-confirms-first-coronavirus-cases-including-first-death/>

^39^ <https://cv.nmhealth.org/2020/03/11/new-mexico-announces-first-presumptive-positive-covid-19-cases/>

^40^ <https://www.youtube.com/watch?v=G7mzYadMBBg> <https://www.health.nd.gov/news/first-case-novel-coronavirus-confirmed-north-dakota-work-continues-prevent-spread>

^41^ <https://governor.arkansas.gov/news-media/press-releases/governor-hutchinson-confirms-states-first-presumptive-positive-covid-19-cas> <https://www.kait8.com/2020/03/11/governor-first-presumptive-case-coronavirus-arkansas/>

^42^ <https://news.delaware.gov/2020/03/11/public-health-announces-first-presumptive-positive-case-of-coronavirus-in-delaware-resident/> <https://whyy.org/articles/delaware-announces-states-first-coronavirus-case-udel-to-move-classes-online/>

^43^ <https://health.wyo.gov/wyomings-first-coronavirus-disease-2019-case-reported/> <https://health.wyo.gov/new-case-added-to-wyomings-coronavirus-total/>

^44^ <https://www.foxnews.com/health/maine-confirms-first-coronavirus-case> <https://www.sunjournal.com/2020/03/12/janet-mills-says-androscoggin-county-woman-is-states-first-presumptive-case-of-covid-19/>

^45^ <https://gov.alaska.gov/newsroom/2020/03/12/first-case-of-covid-19-confirmed-by-alaska-state-public-health-laboratory-is-an-international-resident/>

^46^ <https://www.wdam.com/2020/03/12/forrest-county-man-is-first-presumptive-coronavirus-case-miss/>

^47^ <https://www.idahostatesman.com/living/health-fitness/article240956606.html> <https://healthandwelfare.idaho.gov/news/update-contact-phd-first-case-novel-coronavirus-reported-south-central-idaho-second-state>

^48^ <https://abc3340.com/news/coronavirus/alabama-identifies-first-case-of-covid-19>

^49^ <https://www.kxlh.com/news/helena-news/governor-bullock-confirms-four-presumptively-positive-coronavirus-cases-in-montana>

^50^ <https://www.journal-news.net/journal-news/first-confirmed-wv-coronavirus-case-in-shepherdstown-wife-to-get/article_3033c813-4675-58a1-b127-d91fc97e1178.html>

**Table S5.** Assumptions in detection time model.

| Assumption | Reason |
| --- | --- |
| Detection systems will be implemented in advance in the community originating the outbreak. | This assumes detection systems are implemented broadly as proposed in^19,21^. It is unlikely that detection systems would be implemented in 100% of communities, but we assume coverage in at least the community of origin to show the benefits if such systems are fully funded and implemented. This can be relaxed with a corresponding increase in average detection time. |
| Multiplex diagnostic tests and algorithms can be developed to detect new pathogens by conserved nucleic acid sequences of known pathogen families. | Such multiplex testing may not catch *completely* novel pathogens, but this approach is applicable to most recent emerging pathogens such as SARS-CoV-2 (2019), Ebola (2013), MERS-CoV (2012), and pandemic flu (2009). Proposed technologies include multiplex PCR^41–44^, CRISPR-based multiplex diagnostics^45^, and metagenomic sequencing^46^. Novel pathogens from multiplex testing can be distinguished from known pathogens by sequencing, but one can also apply the model to calculate detection times of new outbreaks of known pathogens. |
| Detection systems are generally modeled assuming that their implemention follows the details in various proposals. | In hospital monitoring, hospitals would test for high-priority pathogen families (e.g. coronaviruses) in patients presenting with severe infectious symptoms in hospital emergency departments^19^. Similarly, in wastewater monitoring, governments would test for pathogens in city wastewater treatment plants daily, and monitor for high and increasing levels of high-priority pathogen families^21^. In air travel monitoring, we model testing of individual symptomatic passengers (differs from proposals to monitor airplane sewage^22^ or bridge air) on incoming international flights for the same pathogens. |
| Status quo detection is modeled as a partially implemented form of hospital monitoring (lower detection probability per case p_test_). | Many recent outbreaks have been detected in healthcare settings^59,69,71,72^. |
